# Objective olfactory testing in patients presenting with sudden onset olfactory dysfunction as the first manifestation of confirmed COVID-19 infection

**DOI:** 10.1101/2020.04.15.20066472

**Authors:** Jerome R. Lechien, Pierre Cabaraux, Carlos M. Chiesa-Estomba, Mohamad Khalife, Jan Plzak, Stéphane Hans, Delphine Martiny, Christian Calvo-Henriquez, Claire Hopkins, Sven Saussez

**Affiliations:** COVID-19 Task Force of the Young-Otolaryngologists of the International Federations of Oto-rhino-laryngological Societies (YO-IFOS); Department of Human Anatomy and Experimental Oncology, Faculty of Medicine UMONS Research Institute for Health Sciences and Technology, University of Mons (UMons), Mons, Belgium; Department of Surgery, Foch Hospital, School of Medicine, UFR Simone Veil, Université Versailles Saint-Quentin-en-Yvelines (Paris Saclay University), Paris, France; Department of Otorhinolaryngology and Head and Neck Surgery, CHU de Bruxelles, CHU Saint-Pierre, School of Medicine, Université Libre de Bruxelles, Brussels, Belgium; Department of Medicine, Neurology, CHU de Charleroi, Charleroi, Belgium; Department of Otorhinolaryngology-Head & Neck Surgery, Hospital Universitario Donostia, San Sebastian, Spain; Department of Head and neck Surgery, EpiCURA Hospital, Hornu, Belgium; Department of Otolaryngology – Head & Neck Surgery, 1st Faculty of Medicine, University Hospital Motol, Prague, Czech Republic; Department of Microbiology, Laboratoire Hospitalier Universitaire de Bruxelles - Universitair Laboratorium Brussel (LHUB-ULB), Brussels, Belgium; Faculté de Médecine et Pharmacie, Université de Mons (UMONS), Mons, Belgium; Department of otolaryngology-Hospital Complex of Santiago de Compostela, Santiago de Compostela, Spain; Guy’s and St Thomas NHS Foundation Trust, London, UK; British Rhinological Society (President), London, UK

**Author notes:** **Correspondence to:** Prof. Sven Saussez, M.D, Ph.D., Department of Human Anatomy and Experimental Oncology, Faculty of Medicine, UMONS Research Institute for Health Sciences and Technology, University of Mons (UMons), Mons, Belgium. Dr Lechien & Dr Cabaraux have equally contributed to this work and should be regarded as *joint first authors*. **Compliance with Ethical Standards**. **Disclosure of potential conflicts of interest:** Authors have no conflict of interest. **Research involving human participants and/or animals:** The ethics committee of Jules Bordet Institute approved the study protocol (IJB-0M011-3137). **Informed consent:** Patients were invited to participate and the informed consent was obtained Conflict of interest statement: The authors have no conflicts of interest.

**Keywords:** COVID-19, Olfactory dysfunction, anosmia, RT-PCR, psychophysical olfactory evaluation

## Abstract

**Background:** The aims of this study are to investigate the COVID-19 status of patients with initial sudden olfactory anosmia (ISOA) using nasopharyngeal swabs for RT-PCR analysis and to explore their olfactory dysfunctions with psychophysical olfactory evaluation.

**Methodology:** This prospective study included 78 ISOA patients who fulfilled a patient-reported outcome questionnaire and underwent a nasopharyngeal swabs. Among these, 46 patients performed psychophysical olfactory evaluation using sniffing tests. Based on the duration of the ISOA, two groups of patients were compared: patients with anosmia duration ≤12 days (group 1) and those with duration >12 days (group 2).

**Results:** Among group 1, 42 patients (87.5%) had a positive viral load regarding RT-PCR while 6 patients (12.5%) were negative. In group 2, 7 patients (23%) had a positive viral load and 23 patients (77%) were negative. Among the 46 patients having performed a psychophysical olfactory evaluation, we observed anosmia in 52% (N=24), hyposmia in 24% (N=11) and normosmia in 24% (N=11) of patients. The viral load significantly decreased throughout the 14-days following the onset of the olfactory disorder.

**Conclusions:** Our results support that a high proportion of ISOA patients are Covid+. Our study supports the need to add anosmia to the list of symptoms used in screening tools for possible COVID-19 infection.

## INTRODUCTION

Since the first case of pneumonia related to severe acute respiratory syndrome coronavirus 2 (SARS-CoV-2), [1] the coronavirus disease 2019 (Covid-2019) has spread rapidly worldwide. The first European cases have been identified in Italy in January, 19, 2020 [2]. As of April 4, a total of 237,544 European patients have been diagnosed through laboratory testing and 25,616 people died from the Covid-19 infection [3]. Anecdotal observations have been rapidly accumulating from many European virologists and otolaryngologists that sudden anosmia and dysgeusia are peculiar symptoms associated with the Covid-19 infection. Hopkins & Kumar published a letter on behalf of the British Rhinological Society describing

“*the loss of sense of smell as a marker of COVID-19 infection*.” They proposed that adults presenting with anosmia but no many other symptoms should self-isolated for seven days [4]. This has been followed by a larger series of 2,428 patients presenting with new onset anosmia during the COVID-19 pandemic of whom 16% reported loss of sense of smell as isolated symptom. The main limitation of this series was the lack of psychophysical testing to confirm the Covid-19 status of the patients [5].

In this context, the Young Otolaryngologists Group of the International Federation of Otorhinolaryngological Societies (YO-IFOS) conducted the first prospective epidemiological study investigating the prevalence of smell and taste disorders. Then, authors reported that 85.6% and 88.0% of mild-to-moderate covid-19 patients reported olfactory and gustatory dysfunctions, respectively[6]. Interestingly, the olfactory dysfunction appeared before (11.8%), after (65.4%) or at the same time (22.8%) as the appearance of general or otolaryngological symptoms. Females were significantly more frequently affected by olfactory and gustatory dysfunction than males [6].

As many patients in the studies by Lechien *et al*. and Hopkins *et al*. developed other symptoms after reporting loss of smell, we wanted to further study all patients with initial sudden onset anosmia, with or without later symptoms, highlighting its importance as one of the first COVID-19 manifestation. We have therefore used the term ‘Initial Sudden Onset Anosmia’ (ISOA) in place of the ‘Isolated’ that was previously described [4].

The aim of this study is to investigate the COVID-19 status of these patients using nasopharyngeal swabs for RT-PCR analysis and to further explore their olfactory dysfunction with objective tests.

## MATERIELS AND METHODS

The ethics committee of Jules Bordet Institute approved the study protocol (IJB-0M0113137). Patients were invited to participate and the informed consent was obtained.

### Subjects and Setting

The clinical data of patients with sudden olfactory dysfunction as first covid-19 manifestation were collected using a questionnaire detailed below. The following inclusion criteria were considered: adult (>18 yo); native French-speaker patients and patients clinically able to fulfill the questionnaire. The following exclusion criteria were considered: patients with olfactory or gustatory dysfunctions before the COVID-19 epidemic; patients with history of chronic rhinosinusitis or nasal polyposis; history of nasal surgery (including rhino/septoplasty with or without functional endoscopic sinus surgery), pregnant woman. We defined our population as having an onset of the anosmia within 15 days (referring to a study demonstrating that the viral load was absent after 15 days) [7] of initial assessment and the lack of general non-otolaryngological symptoms. We further divided our population in two groups: group 1 corresponding to patients with a duration of anosmia ≤12 days and group 2 with patients with a duration of anosmia >12 days. The choice of 12 days is based on the work of Zou *et al*. who they showed that almost all COVID-19+ patients were tested negative 13 days after the onset of the symptoms [7].

### Clinical Outcomes

The online questionnaire was created with Professional Survey Monkey® (San Mateo, California, USA), so that each participant could complete the survey only once. The selection of the relevant epidemiological and clinical features composing the questionnaire was carried out by the Covid-19 Task Force of YO-IFOS, which includes otolaryngologists from North America, Europe and Asia [8]. The demographic and clinical outcomes consisted of the assessment of nasal obstruction, rhinorrhea, postnasal drip, throat pain, facial pain, ear pain, dysphagia and dysgeusia, the latter being defined as the impairment of the following four taste modalities: salty, sweet, bitter and sour.

### Olfactory & Gustatory Outcomes

The olfactory and gustatory questions were based on questions from the smell and taste component of the National Health and Nutrition Examination Survey [9]. This population survey was implemented by the Centers for Disease Control and Prevention to continuously monitor the health of adult citizens in the U.S. through a nationally representative sample of 5,000 persons yearly [9]. The questions have been chosen to characterize the variation, timing and associated-symptoms of both olfactory and gustatory dysfunctions, and, therefore, they suggest a potential etiology.

### Nasopharyngeal swabs for RT-PCR

A nasopharyngeal swab was performed by two senior otolaryngologists (MK and SS). Specimens were immediately sent to the laboratory LHUB-ULB (Laboratoire Hospitalier Universitaire Bruxelles - Universitair Laboratorium Brussel), Brussels. The microbiological confirmation of SARS-CoV-2 was performed by reverse transcription polymerase chain reaction (RT-PCR) assays. Viral RNA extraction was performed by m2000 mSample Preparation SystemDNA Kit (Abbott) using 1000µl manually lysed sample (700µl sample + 800µl lysis buffer from kit) eluted in 90µl elution buffer. A qRT-PCR internal control was added at each extraction. qRT-PCR was performed using 10µl of extracted sample in the RealStar^®^SARS-CoV-2 RT-PCR Kit from altona-diagnostics with a cut-off set at 40 Ct.

### Psychophysical Olfactory Evaluation

We used the Identification Test of the ‘Sniffin Sticks’ test (Medisense, Groningen, The Netherlands) to evaluate olfactory performance in ISOA patients. All patients were invited to attend for testing. Sniffin Sticks is a fully validated test for objective testing of olfactory disturbance [10]. A total of 16 pens were presented to the patient at 30 second of intervals. The patient had to choose the term which describes the presented odorant best from 4 given options and has to make a choice, even if unsure. The test was scored on a total of 16 points and allowed categorization into in 3 groups: normosmia (score between 12-16), hyposmia (score between 9-11) and anosmia (score 8 or below). Tests were performed on the same day as completion of patient rated olfactory and gustatory outcomes.

### Statistical Analyses

Statistical analyses were performed using the Statistical Package for the Social Sciences for Windows (SPSS version 22,0; IBM Corp, Armonk, NY, USA). The relationship between CT (inversely reflecting viral load) and the duration of olfactory dysfunction was assessed by Spearman rho test. A level of significance of p<0.05 was used.

## RESULTS

A total of 78 patients were identified with sudden olfactory dysfunction as the first manifestation of possible COVID-19 infection. Table 1 describes demographic and epidemiological characteristics of patients. The mean age of patients was 40.6 ± 11.2 years old (21–67). There were 46 (59%) females and 32 (41%) males. The following ethnicities composed the cohort: Caucasian 73 (93.6%), North African 4 (5.1%) and Sino-African 1 (1.3%). The most prevalent comorbidities of patients were gastroesophageal reflux disease (GERD) (11.5%), allergic rhinitis (9%), asthma (5%) and hypertension (5%). 86% of patients are non-smokers. There was no patient with chronic rhinosinusitis.

**Table 1:**
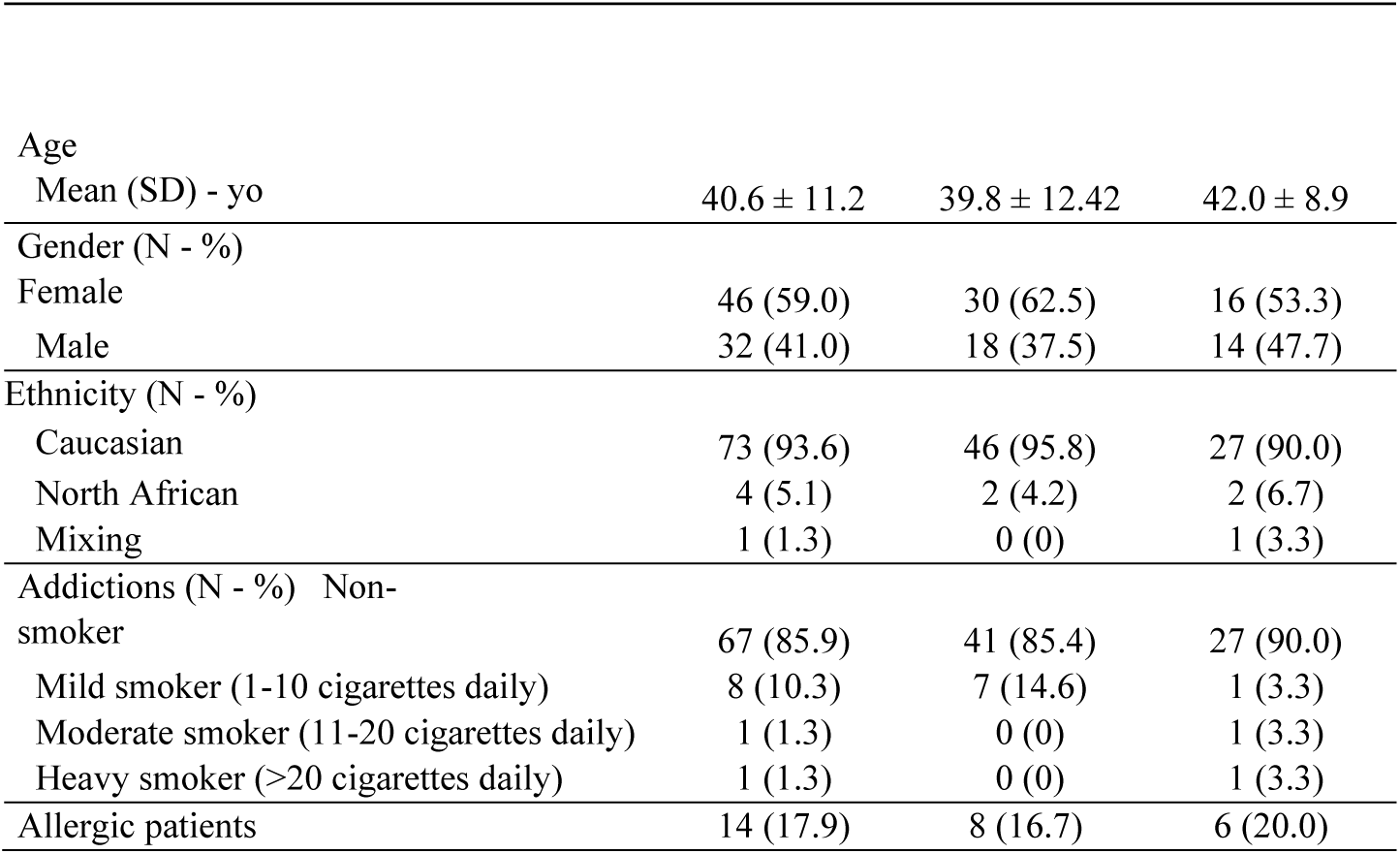
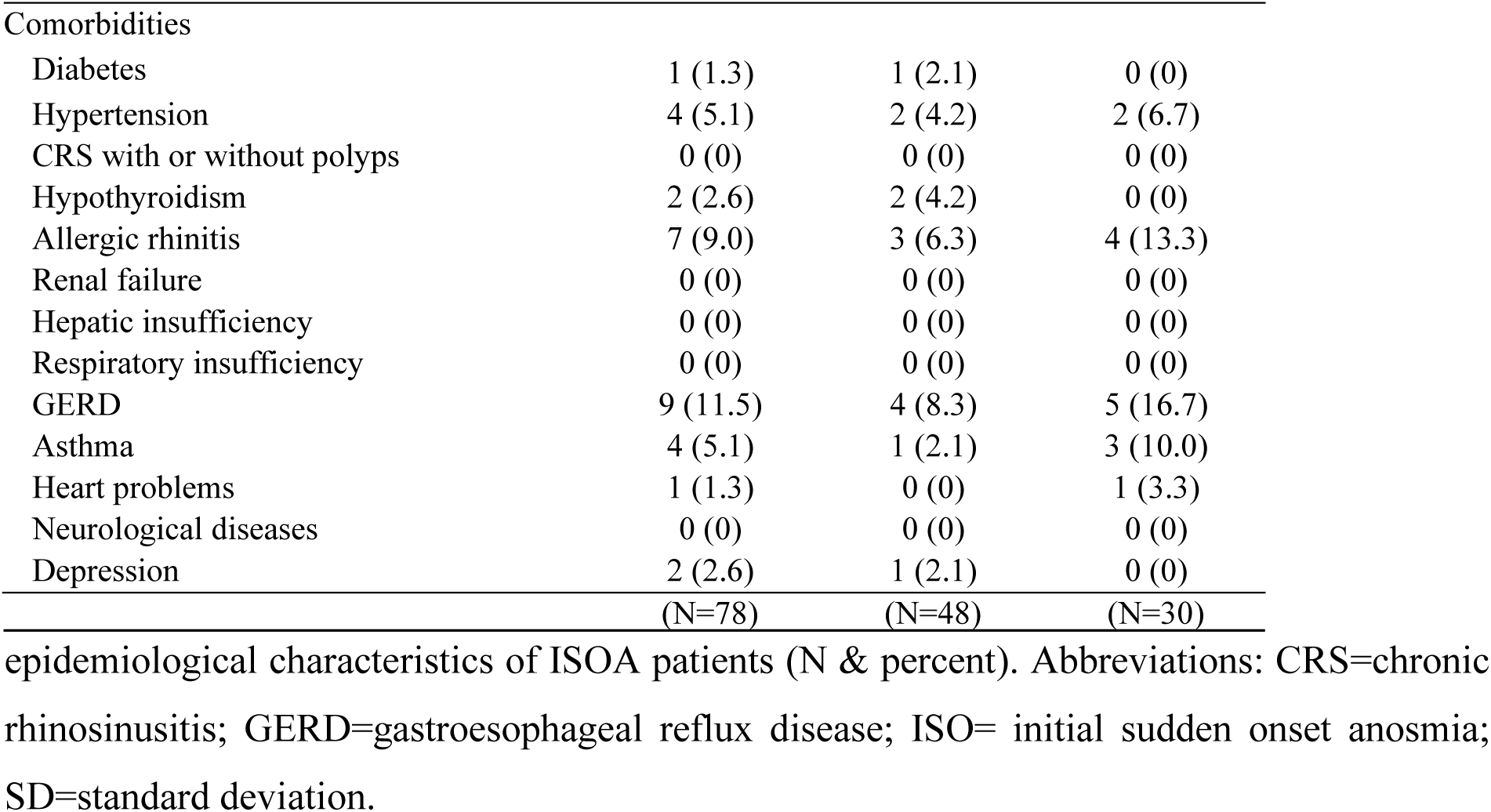
demographic and epidemiological characteristics of ISOA patients.

### Clinical Outcomes

The otolaryngological complaints are described in Table 2. The most prevalent otolaryngological symptoms during the clinical course of the disease were: dysgeusia (67.9%), postnasal drip 46.2%) and nasal obstruction (46.2%). A total of 35 patients (44.9%) presented sudden olfactory dysfunction without nasal obstruction or rhinorrhea. Cacosmia and phantosmia were reported during the clinical course by 57 and 38 patients, respectively. The aroma perception was reduced (N=18), disappeared (N=29) or distorted (N=5) in 23.1%, 37.2%, and 6.4%, respectively. 52.7% of patients reported that the loss of smell was constant and did not change at the onset of the disease.

**Table 2:** Ear, nose & throat symptoms in ISOA patients ISOA patients Ear, nose & throat symptoms Table 2 describes the ear, nose and throat symptoms (N & percent) of ISOA patients.

|  |  |  |  |
| --- | --- | --- | --- |
| Nasal obstruction | 38 (48.7) | 25 (52.1) | 13 (43.3) |
| Rhinorrhea | 31 (39.7) | 19 (39.6) | 12 (40.0) |
| Postnasal drip | 36 (46.2) | 22 (45.8) | 14 (46.7) |
| Throat pain | 23 (29.5) | 15 (31.3) | 8 (26.7) |
| Facial pain | 26 (33.3) | 15 (31.3) | 11 (36.7) |
| Ear pain | 19 (24.4) | 11 (22.9) | 8 (26.7) |
| Dysphagia | 21 (26.9) | 13 (27.1) | 8 (26.7) |
| Dysgeusia | 53 (67.9) | 31 (64.6) | 22 (73.3) |

### COVID-19 RT-PCR positivity and ISOA patients

Among group 1 (anosmia ≤12 days), 42 patients (87.5%) had a positive viral load regarding RT-PCR COVID-19 while 6 patients (12.5%) were negative. In group 2 (anosmia >12 days), 7 patients (23%) had a positive viral load and 23 patients (77%) were negative.

We analyzed the viral load in nasopharyngeal swabs obtained from our ISOA patients in relation to day of onset of any symptoms. Figure 1 shows that the higher viral loads (inversely related to Ct value) were statistically correlated to shorter ISOA delay (r_s_=0.441, p=0.004).

**Figure 1:**
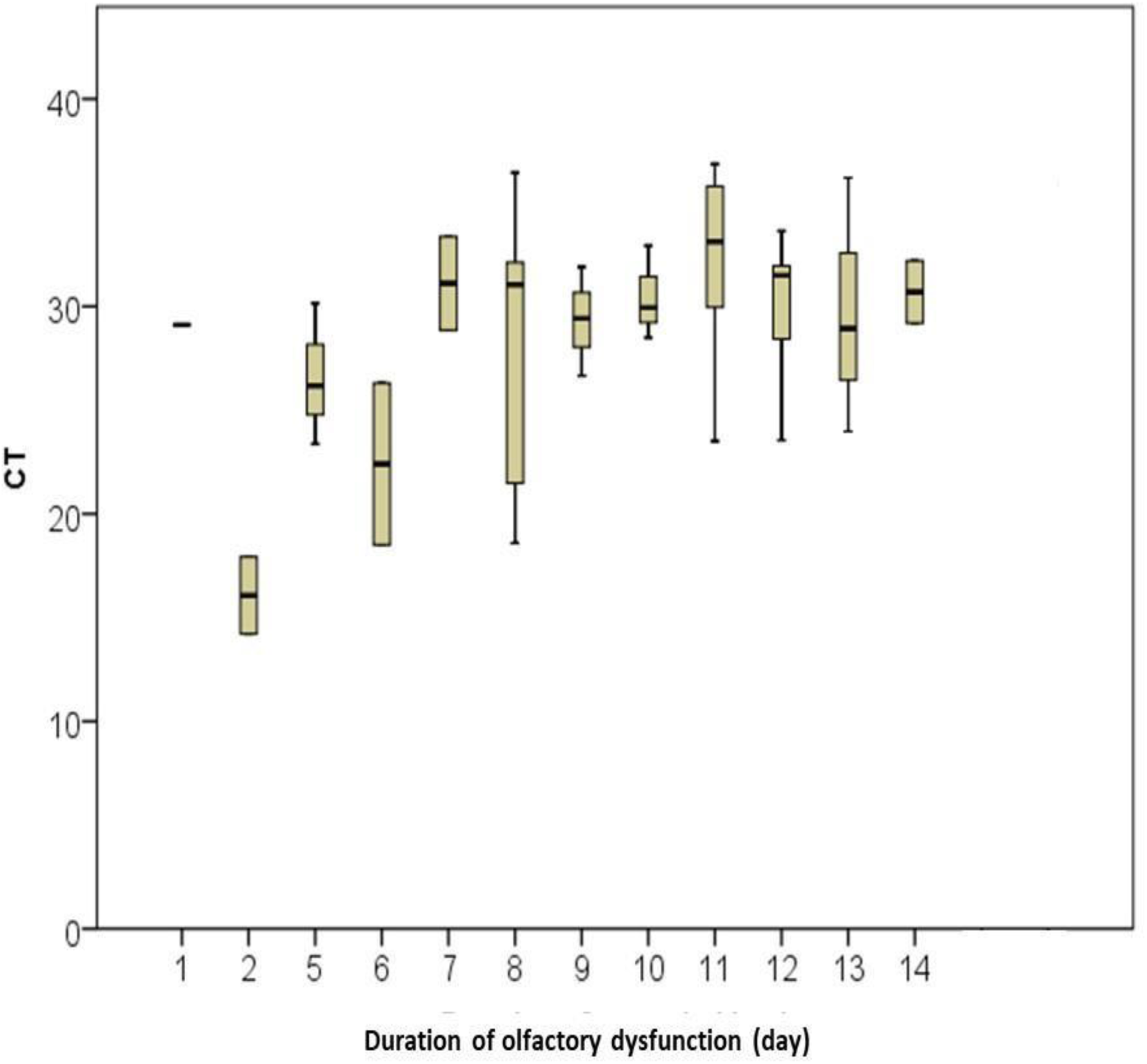
Duration of olfactory dysfunction in relationship with the CT (reflecting inversely the viral load)

### Psychophysical Olfactory Evaluation and ISOA patients

All patients were invited to attend for olfactory testing but due to restrictions on travel and the patient fear of contamination, this could only be performed in 46 patients. Among the 48 (group 1) patients, 21 RT-PCR COVID-19 + patients also performed also a sniffing test. Among these 11 patients (52%) were rated as anosmic, 4 patients (19%) hyposmic and 6 patients normosmic (29%). Among the 4 RT-PCR COVID-19-patients, 2 patients were anosmic, 1 hyposmic and 1 normosmic. For most patients, the sniffing test and the questionnaire were done the same day. Among the 30 (group 2) patients, 7 RT-PCR COVID-19 + patients performing a sniffing test, 3 patients (43%) were anosmic, 3 patients (43%) hyposmic and 1 patients (6%) normosmic. From the 14 RT-PCR COVID-19-patients 8 patients were anosmic (58%), 3 hyposmic (21%) and 3 normosmic (21%).

Reuniting all patients (46 patients) having performed objective olfactory evaluation (sniffing test), we can observed that 24 patients were anosmic (52%), 11 patients hyposmic (24%) and 11 patients normosmic (24%).

## DISCUSSION

Recently, eminent clinicians observed abrupt loss of smell COVID-19 infected individuals, particularly from Britain, the US, France, South Korea, China, Germany, and Iran. They collectively reinforced its potential application as a marker that could be used in the first line of diagnostics in patients presenting with COVID-19 symptoms. The Hopkins team published the first case report and case series suggesting that sudden olfactory dysfunction, should be considered highly suspicious for SARS-CoV-2 [4,6]. Moreover, our study group recently showed, that anosmia was present in 86% in a series of 417 patients with mild-to-moderate COVID-19 disease, and appeared before other symptoms in 11.8% [5]. In the same way, Kaye *et al*. described in a series of 237 patients that anosmia was noted in 73% of subjects prior to COVID-19 diagnosis and was the initial symptom in 26.6% [11]. In a larger series of 1,420 mild to moderate COVID+ patients, our Bayesian analysis identified that reported anosmia was a key symptom in COVID-19 infection with 70.2% of cases (Lechien *et al*., submitted). However, these studies suggest that truly isolated anosmia is uncommon, and many patients have other otolaryngological symptoms.

We therefore set out to study the COVID-19 status in patients presenting with sudden olfactory disturbance as the first manifestation. Our study demonstrates that 87.5% of 48 patients with an anosmia duration ≤12 days were RT-PCR COVID-19+. In this group (anosmia duration ≤12 days), 21 patients have performed a sniffing test, which demonstrated that 71% were anosmic or hyposmic and 29% normosmic. For patients with an anosmia duration >12 days, 7 patients (23%) were RT-PCR COVID-19+ and 23 patients (77%) negative. We think this likely reflects recovery and viral clearance in some of this group. We excluded patients with pre-existing chronic rhinosinusitis or prior nasal surgery from our study and therefore it is likely that the additional otolaryngological symptoms described by almost 50% of our patients are associated with their COVID-19 infection.

This is the first study providing objective smell evaluations in patients reporting loss of sense of smell during the COVID-19 pandemic. Considering all patients who performed a sniffing test, we showed that 52% were anosmic, 24% hyposmic and 24% normosmic. The same trend was observed in our both groups (anosmia ≤12 days *versus* anosmia >12 days) although the expected the proportion who were anosmic decreased slightly to 43% in the group with longer duration

Akerlund *et al*. have studied the olfactory threshold (using a discrimination test with dilution of butanol) and nasal mucosa changes by acoustic rhinometry in experimentally induced common cold after nasal inoculation of coronavirus HCV209 in 20 volunteers [12]. They showed that in individuals with a cold (9 volunteers) and impaired olfaction, the change of smell ability was correlated to nasal obstruction. Based on Akerlund *et al*. study patient reported symptoms suggesting that COVID-19 infection provokes nasal obstruction, we hypothesized that this could explain the anosmia [12]. However, although a significant proportion (49%) of our patients reported nasal obstruction, the majority did not, suggesting neural mechanisms are also involved.

In 2007, Suzuki *et al*. demonstrated that coronavirus may be detected in the nasal discharge of patients with olfactory dysfunction [13], and that some patients had normal acoustic rhinometry, suggesting that nasal inflammation and related obstruction were not the only etiological factors underlying the olfactory dysfunction in viral infection. Netland *et al*. demonstrated on transgenic mice expressing the SARS-CoV receptor (human angiotensinconverting enzyme 2) that SARS-CoV may enter the brain through the olfactory bulb, leading to rapid transneuronal spread [14]. Interestingly, authors demonstrated that the virus antigen was first detected 60 to 66 hours post-infection and was most abundant in the olfactory bulb. Recently, Gupta *et al*. performed a bioinformatic analysis of single-cell expression profiles underscored selective expression of angiotensin coverting enzyme 2 (*ACE2)* in a subset of horizontal basal cells and sustentacular cells of the olfactory mucosa in humans [15]. They evaluated the expression of *ACE2* transcript in 3,906 olfactory mucosa originated single cells from the recent report by Durante *et al*. [16] and suggested that loss of smell in the infected patients is most unlikely due to the direct impairment of the olfactory sensory neurons; in particular the sustentacular cells and the horizontal basal cells are the potential cell types that are highly susceptible to viral entry.

It is interesting to note that 24% of patients were found to have normal olfactory function on testing. It is known that patients sometimes have difficulty rating their own sense of smell (particularly in the setting of associated nasal obstruction), [17] and the objective test results suggesting that some patients actually have normal odor identification ability may reflect this. It may be that olfactory thresholds were reduced but not the ability to identify odorants; future studies should include threshold and discrimination testing (which were not performed due to difficulty in preventing contamination of the testing kits) and simultaneous patient rated and objective tests of olfactory function.

## CONCLUSIONS

The results of this study give further support to the urgent need to add anosmia to the list of symptoms used in screening tools for possible COVID-19 infection. The current evidence base makes it untenable to continue to disregard this symptom any longer. Use of loss of smell -even if occurring in the absence of non-otolaryngological symptoms-as a marker will be a very useful weapon in the COVID-19 fight, especially in countries where access to testing will be greatly limited. Future clinical and basic science researches are needed to better understand the pathophysiological mechanisms underlying the development of olfactory dysfunction in COVID-19 patients.

## Data Availability

All data are available

## ACKNOWLEDGMENTS

The University of Mons head and the “call center” staff coordonnated by Ahmed Cherifi and the following collaborators and researchers: Philippe Boelpaep, Anne Trelcat, Geradine Descamps, Sonia Furguelese, Laura Soumoy et Fabrice Journe.

We thank the University of Mons (UMONS) for funding this work, as well as FRMH grant.

## AUTHORSHIP CONTRIBUTION

S.S and J.L initiated, designed the project and developed the methods. S.S, J.L, C.H wrote the manuscript. J.P, S.H, C.C.E, C.C.H, D.M read, corrected and commented the manuscript. S.S., M.K and P.C performed the experiments. D.M performed the RT-PCR analysis. S.S, J.L and C.H analyzed the results. C.H, J.P, S.H supervised the project. All authors read and approved the final manuscript.

